# Remote Intensive Parent-Implemented Intervention for Young Children on the Autism Spectrum During Covid-19: The Experience of Parents and Therapists

**DOI:** 10.1101/2022.01.30.22270029

**Authors:** Hilary Wood de Wilde, Nada Kojovic, Céline Robertson, Catherine Karr, Leyla Akman, Florence Caccia, Astrid Costes, Morgane Etienne, Martina Franchini, Edouard Gentaz, Marie Schaer

## Abstract

In response to a Covid-19 period of home-confinement, autism early intervention programs in Geneva, Switzerland, converted their in-person services to a telehealth format. Forty-five families received daily videoconferencing sessions of primarily parent-implemented intervention. Questionnaires were completed at three time points. Child progress was monitored using the Early Start Denver Model Curriculum Checklist. Parents maintained high levels of participation and satisfaction, regardless of socio-economic or cultural background, with the majority reporting an improvement in their use of intervention techniques. Child progress followed a pattern of continued significant improvement across most developmental domains. Findings suggest that a more frequent dosage of parent-implemented intervention than typically studied is not only feasible, but appreciated by caregivers, especially when delivered via the time-saving videoconferencing format.

---

In the spring of 2020, with the spread of the Covid-19 pandemic, social distancing measures prevented many families with children on the autism spectrum from accessing in-person early intervention services (Crockett et al., 2020; Ellison et al., 2021; Kronberg et al., 2021). The preschool years represent a critical developmental period in which children who receive evidenced-based intervention can make significant gains in language, cognitive and adaptive functioning (Fuller & Kaiser, 2020; Schreibman et al., 2015). Faced with home-confinement orders and cancelled programs, many caregivers and professionals looked for alternative ways to maintain intensive early intervention instruction (Crockett et al., 2020; Ellison et al., 2021; Kronberg et al., 2021). The current article reports on 45 families in Geneva, Switzerland who replaced their child’s in-person intervention program with remote telehealth sessions for a two-month period.

The majority of sessions provided were parent implemented intervention (PII; also referred to as ‘parent coaching’), where a caregiver is coached to use techniques that foster engagement and promote a child’s learning (Oono et al., 2013; Rogers, Estes, et al., 2012). Research has shown PII to be a valuable part of early intervention in autism (Green et al., 2010; Oono et al., 2013; Rogers, Estes, et al., 2012; Zwaigenbaum et al., 2015). Coaching parents remotely via telehealth has also proven feasible, a cost-effective way to increase access to high quality services (de Nocker & Toolan, 2021; Ellison et al., 2021; Ingersoll & Berger, 2015; Vismara et al., 2018).

The effect of either in-person or remote PII has usually been studied at the dosage of one to three hours per week (de Nocker & Toolan, 2021; Oono et al., 2013; Parsons et al., 2017; Rogers et al., 2019). This frequency appears to be arbitrarily determined, based perhaps on the funding available, or on an estimation of how often caregivers would agree to participate in sessions, or of how much time they required between sessions to consolidate their learning. Recent research, however, suggests that parents and their children may benefit from more frequent PII sessions (Rogers et al., 2019; Wetherby et al., 2014). In order to define an optimal frequency, one in which the caregiver manages to acquire the intervention skills quickly while still maintaining a high level of overall satisfaction, there needs to be a better understanding of how families of young autistic children, and the professionals working with them, experience a higher dosage of PII sessions. The current study examines the engagement and satisfaction of parents and therapists as they experienced a substantial increase in parent coaching sessions provided via videoconference.

Importantly, these parents represent a diverse population. Research has, until recently, tended to study parent coaching with volunteers who were typically white, college-educated mothers, and not employed outside the home (Ellison et al., 2021; Ingersoll & Berger, 2015; Vismara et al., 2018). Less is known about how working parents, fathers, families with lower socioeconomic status or families from diverse cultural backgrounds experience these autism services (Mirenda et al., 2022; Rogers et al., 2022; Stahmer et al., 2019). This has meant that the majority of caregivers who could potentially benefit from PII services are not well represented in the literature (Ingersoll & Berger, 2015; Stahmer et al., 2019). Researchers have begun to look at how evidence-based practices, proven to work in clinical autism research settings, can be adapted to work in diverse community settings (Mirenda et al., 2022; Nahmias et al., 2019; Rogers et al., 2022; Stahmer et al., 2019). The Covid-19 home-confinement period in Geneva created a situation in which all families from community-based autism intervention programs received services via videoconference, enabling us to assess the feasibility of providing intense, remote parent coaching to families from many walks of life.

## Methods

### Context of Covid-19 Home Confinement in Switzerland

On the 16^th^ of March, 2020, in response to the initial outbreak of the Covid-19 pandemic, the Swiss Federal Office of Public Health ordered a nationwide closure of all non-medical services, including schools and childcare centers. Our intervention centers re-opened gradually starting April 27th, 2020 with schools re-opening May 11^th^, 2020 (figure 1). Families were given the option to continue sessions online through to the end of May, and despite subsequent social distancing measures, schools and intervention centers have remained open to date.

**Figure 1.**
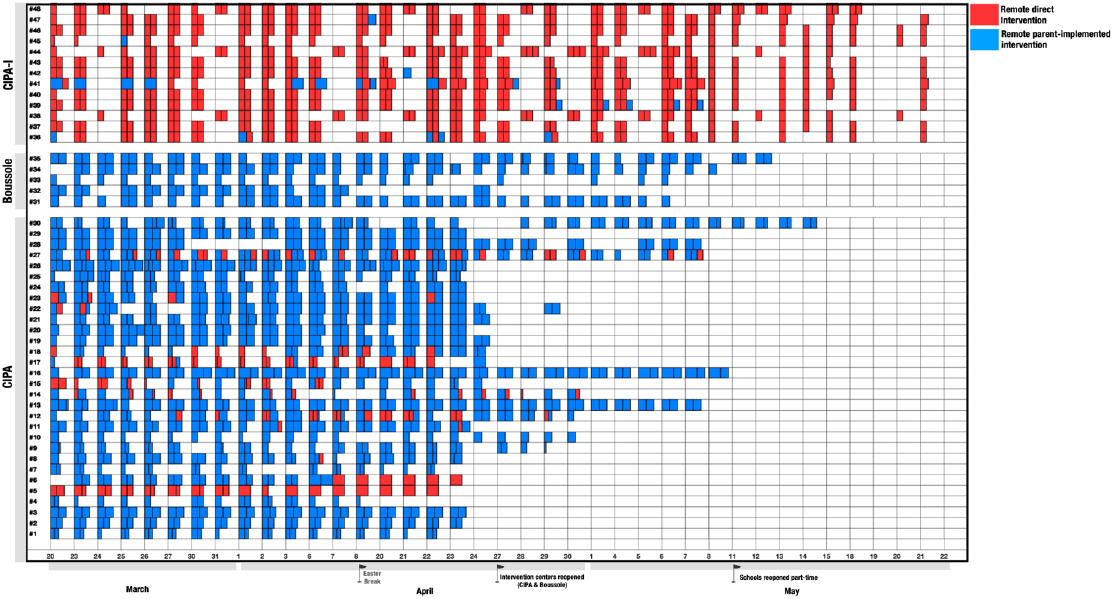
Remote Therapy Sessions Distribution *Note*. The three centers are on the Y axis: CIPA, n = 30; Boussole, n = 5; CIPA-I, n = 13, with each row representing sessions for one child. The timeline is on the X axis. Individual sessions are depicted in colored rectangles (blue = PII and red = DI). Session duration (30, 60, or 90 minutes) is proportional to the width of the rectangles.

### Participants

#### Children

All 48 children enrolled in three intensive intervention programs at the time of the home-confinement were offered remote services (table 1). These participants were part of an already established longitudinal study on the developmental trajectories of young autistic children (Robain et al., 2020). Forty-three children had a diagnosis of Autism Spectrum Disorder (ASD) according to the DSM-5 (American Psychiatric Association, 2013), confirmed using a full-battery evaluation. Five toddlers were suspected of having autism but did not yet have a confirmed diagnosis.

**Table 1.**
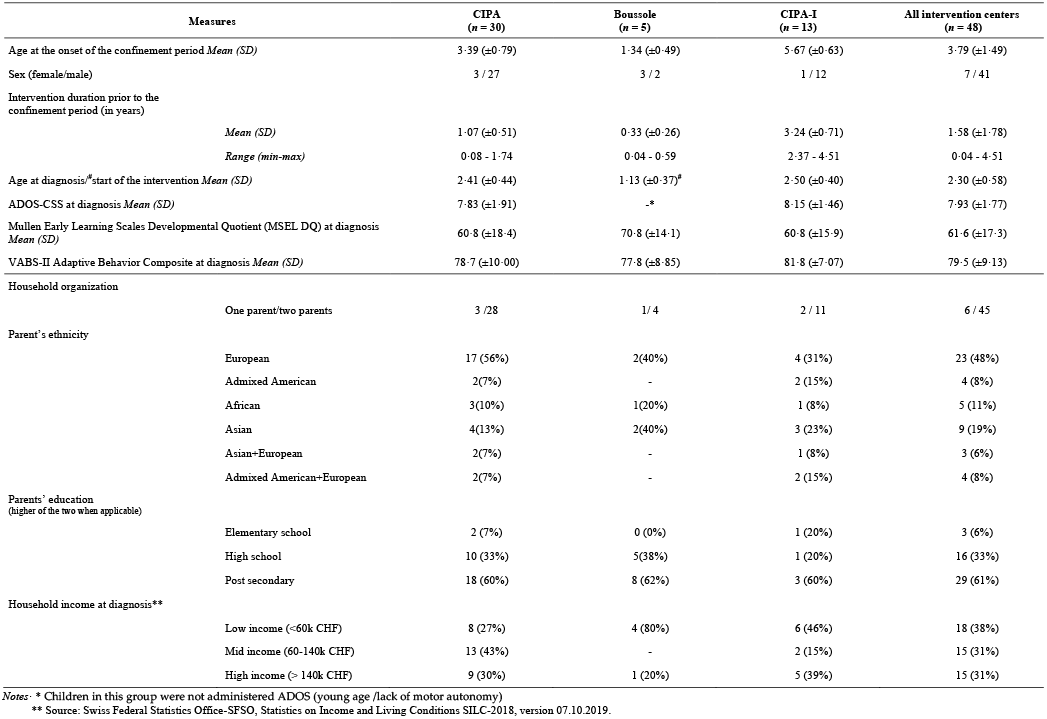
Description of the Sample of Children (n = 48) and Their Families (n = 45)

#### Caregivers

The 45 families (three families had siblings enrolled) had varying socioeconomic status levels, with 38% considered low-income as indexed by the cost-of-living standards in Switzerland (table 1). Educational attainment also varied, with 39% having secondary school or less. Five children came from single-parent households. Parents came from 29 different countries of origin and spoke 15 different languages at home, representative of Geneva’s multicultural demographic (table 2). All families provided their written consent for the study, in accordance with protocols approved by the institutional review board of the University of Geneva.

**Table 2.**
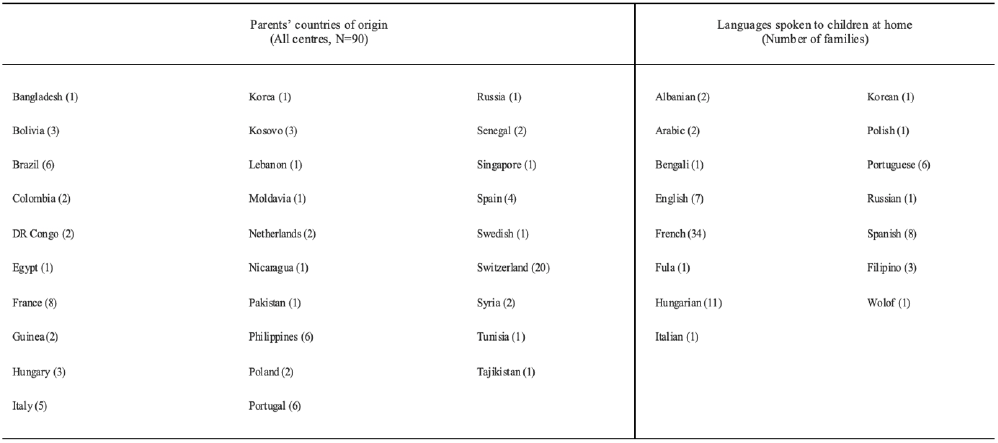
List of Parent’s Countries of Origin and Languages Spoken to Children at Home

#### Therapists

All 45 early intervention therapists working in the intervention programs at the time of the home-confinement, participated in the study. They were all female, aged between 25 and 38 years old.

### Intervention received prior to home-confinement: Three programs

#### 1. Centre for Early Intervention in Autism (Centre d’Intervention Précoce en Autisme / CIPA)

Thirty children aged 24 to 54 months received 20 hours a week of in-center, therapist-delivered intervention services using the Early Start Denver Model (ESDM) (Rogers & Dawson, 2010b). ESDM is an evidenced-based Naturalistic, Developmental Behavioral Intervention (NDBI) for young children on the autism spectrum (Schreibman et al., 2015). In addition, parents initially receive 12 sessions of once-a-week, in-person parent coaching using the Parent-Implemented Early Start Denver Model (P-ESDM). P-ESDM is an effective PII intervention aimed at coaching caregivers as they learn to embed social-communicative learning opportunities for their child into daily routines (Rogers, Estes, et al., 2012). Parents met with their child’s lead therapist one to two times a month, as needed, following the initial P-ESDM sessions. The CIPA program is subsidized by State, private foundation and federal disability insurance funds, and available to children living in the Canton of Geneva. Children typically remain in the CIPA for two years (see table 1 for intervention duration prior to confinement period).

#### 2. The Boussole Program

Five young children suspected of having autism, aged 8 to 24 months, were receiving eight hours a week of in-home and in-center, therapist-delivered ESDM intervention, and two hours of P-ESDM coaching. The Boussole is subsidized by private foundation and insurance funds.

#### 3. The CIPA Inclusion Program (CIPA-I)

Thirteen children aged 4.7 to 6.6 years were receiving 13 to 24 hours of one-to-one in-school intervention services per week. They had all completed the CIPA program prior to starting school. Seven were in their first year of school and six in their second year. The CIPA-I program is provided for two years, and is funded by Geneva’s Department of Public Education.

### Telehealth Intervention During Covid-19 Home Confinement

Each child’s program was individualized for remote delivery, taking into account parameters such as the parents’ availability, their level of access to technology, and the child’s likelihood to attend to a therapist via screen. The families were offered two 1-hour sessions a day, Monday to Friday, of either: 1) Remote Parent-Implemented Intervention (PII), where the therapist provides parent coaching using the P-ESDM approach (Rogers, Estes, et al., 2012), or 2) Remote Direct Intervention (DI) between child and therapist, where the therapist works on the child’s learning objectives using ESDM strategies, proposing interactive activities like singing songs, discussing topics and playing turn-taking games.

### Therapist Training

All therapists were licensed psychologists with a minimum of a master’s degree. They had been trained in the use in of the ESDM approach, meeting fidelity in clinical practice on the ESDM Fidelity Rating System (Rogers & Dawson, 2010b) prior to the home confinement period. Within the team, 15 credentialed ESDM therapists supervised the children’s programs, overseen by an ESDM certified trainer. Therapists had no prior experience in telehealth service provision, and were provided with a one-day emergency training prior to their first session. The majority of sessions were recorded for supervisory purposes and reviewed during remote weekly team clinic meetings.

### Materials

ZOOM software (ZOOM Video Communications, Inc, Version 4·4; https://zoom.us/) was used for videoconferencing sessions. Parents were asked to set up their computer, smartphone or tablet in a place where the therapist would be able to have a clear view of a play area or of the child’s work area. Prior to starting, lead therapists contacted the parents to explain how to install the ZOOM software and to discuss materials needed, such as toys, felt markers and paper. A toy-lending program was arranged for families in need. The book *An Early Start for Your Child with Autism: Using Everyday Activities to Help Kids Connect, Communicate, and Learn* (Rogers, Dawson, et al., 2012) (French, English or other translations) was recommended to families. When appropriate, the therapists also suggested videos, website links and ideas for activities in line with each child’s intervention objectives.

### Measures

#### Parent and Therapist Satisfaction Questionnaires

Satisfaction questionnaires were sent via e-mail to both parents and therapists after 1) the first remote intervention session, 2) at 3 weeks and, 3) after their final online session. Participants responded to questions on a Likert seven-point scale ranging from “Strongly Disagree” to “Strongly Agree” and multiple-choice questions, and were given the opportunity to add written comments pertaining to their experience with the services.

#### The Early Start Denver Model Curriculum Checklist for Young Children with Autism (ESDM-CC)

For this study, the ESDM-CC (Rogers & Dawson, 2010a) was used to monitor developmental progress in the children’s learning objectives, in light of the drastic change to our service delivery model. The children in the Boussole and the CIPA are typically evaluated using the ESDM-CC every 12 weeks. This evaluation helps parents and therapists decide which learning targets they will prioritize in the child’s treatment plan over the following three-month period. The ESDM-CC evaluates areas of expected child development at 4 levels: 9-18 months, 18-24 months, 24-36 months and 36-48 months, covering domains such as communication, social skills, motor skills and cognition (Rogers & Dawson, 2010b). In the current study, we looked specifically at domains that span all four levels: receptive and expressive language, social skills (including joint attention in level 2), and play. Skills are evaluated as being “A” (acquired, consistent, and generalized); “P” (Partial or emerging); “N” (child is unable or unwilling to perform the skill); or “X” (no opportunity or not tested). We considered a child’s current developmental age range to be where the majority (50% or more) of the skill items were rated as an “A”. If the highest level had an equal number of acquired and not-yet acquired skills, the child was considered between two levels for that skill (i.e., 1 / 2), and were converted into numerical levels for the purpose of the statistical analyses (1 / 2 = 1.5; 2 /3 = 2.5, etc.). To compare the magnitude of progress during in-person intervention sessions, we considered three time points: T1- ESDM-CC three-months prior to the Pre-Confinement ESDM-CC; T2 - Pre-Confinement ESDM-CC; T3 - Post-Confinement ESDM-CC.

### Statistical Analyses

We used the Mann Whitney test to examine group differences in program satisfaction, as the measures did not follow a normal distribution (Shapiro-Wilk test). We compared the parents’ and therapists’ overall experience and satisfaction with the remote intervention, as well as the change in satisfaction between the first day and the third week of treatment. To illustrate child developmental progress, we used the Wilcoxon matched-pairs signed-ranks test to compare the global and subdomain scores of the ESDM-CC between the three time points, as the measures were not normally distributed. Statistical analyses were conducted using GraphPad Prism v.8 and SPSS v.25 software (https://www.ibm.com/analytics/spss-statistics-software). All results in our study were considered significant at the p value of 0.05. The False Discovery Rate (FDR) was controlled using Benjamini-Hochberg procedure (Benjamini & Hochberg, 1995).

## Results

### Program Participation

All 48 children and their 45 families, as well the 45 therapists of the CIPA, Boussole and CIPA-I programs, participated in the remote sessions during the confinement period (figure 1; table 3). Thirty-four of the 35 children from the CIPA and Boussole programs received Remote Parent-Implemented Intervention (PII), of which 11 also had some sessions of Remote Direct Intervention (DI). One older child in the CIPA (3 years, 9 months), and all 13 of the children in the CIPA-I program received only DI, although their parents were present in the background. Therapists adapted the length of sessions to the needs and availability of the families, with sessions lasting between 30 and 90 minutes (figure 1).

**Table 3.**
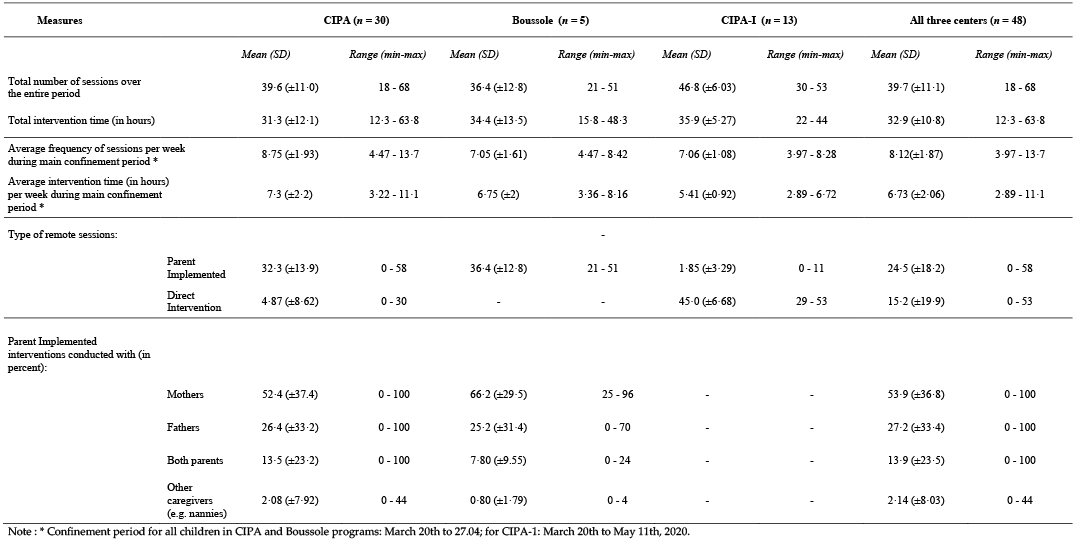
Description of the Remote Intervention Sessions

Remote sessions were offered over a period of 2 months, with a seven-workday break over the Easter holiday (figure 1). During the time when all families were in home-confinement, services were provided for a total of 19 days for the families in the CIPA and Boussole (between March 20^th^ and April 27^th^, 2020) and 29 days for the CIPA-I (between March 20^th^ and May 11^th^, 2020). Children in the CIPA received an average of 8.75 sessions (7.3 (±2.2) hours) of primarily PII sessions per week. The children in the CIPA-I received an average of 7.06 sessions (5.41 (±0.92) hours) of DI per week (table 3).

Thirty-four families, including 56 individual parents, participated in the PII sessions. Twenty-seven percent of the PII sessions were done with fathers, 54% with mothers, 14% with both mother and father, and 2 % with a childcare provider (table 3). Fifty-nine percent of the families used a computer, 24% a tablet, while 16% participated via smartphone. One family did not have access to a wi-fi connection and was coached via landline telephone.

### Parent and Therapist Satisfaction

Forty-two of the 45 families and all 45 therapists filled out all three time-points of the satisfaction questionnaires. Parent(s) who participated in the coaching or were present during the DI sessions completed the questionnaires. We divided the questionnaire responses into the following four areas of interest (figures 2-5).

**Figure 2.**
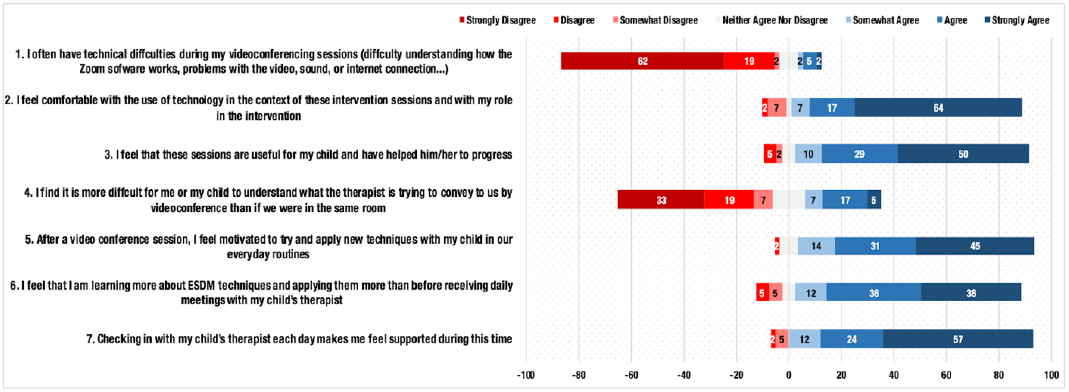
Parents’ Satisfaction with Remote Intervention Delivery Assessed at Week 3 *Note*. Questions were assessed on a 7-point Likert scale (Strongly Disagree-Strongly agree). The percentages of point responses are represented with the exception of the neutral point 4, *Neither agree nor disagree*.

#### 1. Experience and Satisfaction with Online Format

Parents reported general satisfaction with the remote format, with 88% saying they felt comfortable with the technology and their role in the telehealth intervention, and only 9% reporting technical difficulties (figure 2). While 29% found it was harder to understand the therapist via videoconference, nearly all parents (90%) felt motivated to try and apply new techniques with their child following a remote session, and 89% felt the sessions were useful and helped their child to progress. At the end of the home-confinement period, when asked what they found most challenging, 50.3% of parents said it was controlling the home environment around them during the sessions (i.e., siblings and other distractions), while 33% felt it was difficult to manage their own child’s behaviors. Only one parent found it difficult to play with their child while being observed remotely (figure 3, A1).

**Figure 3.**
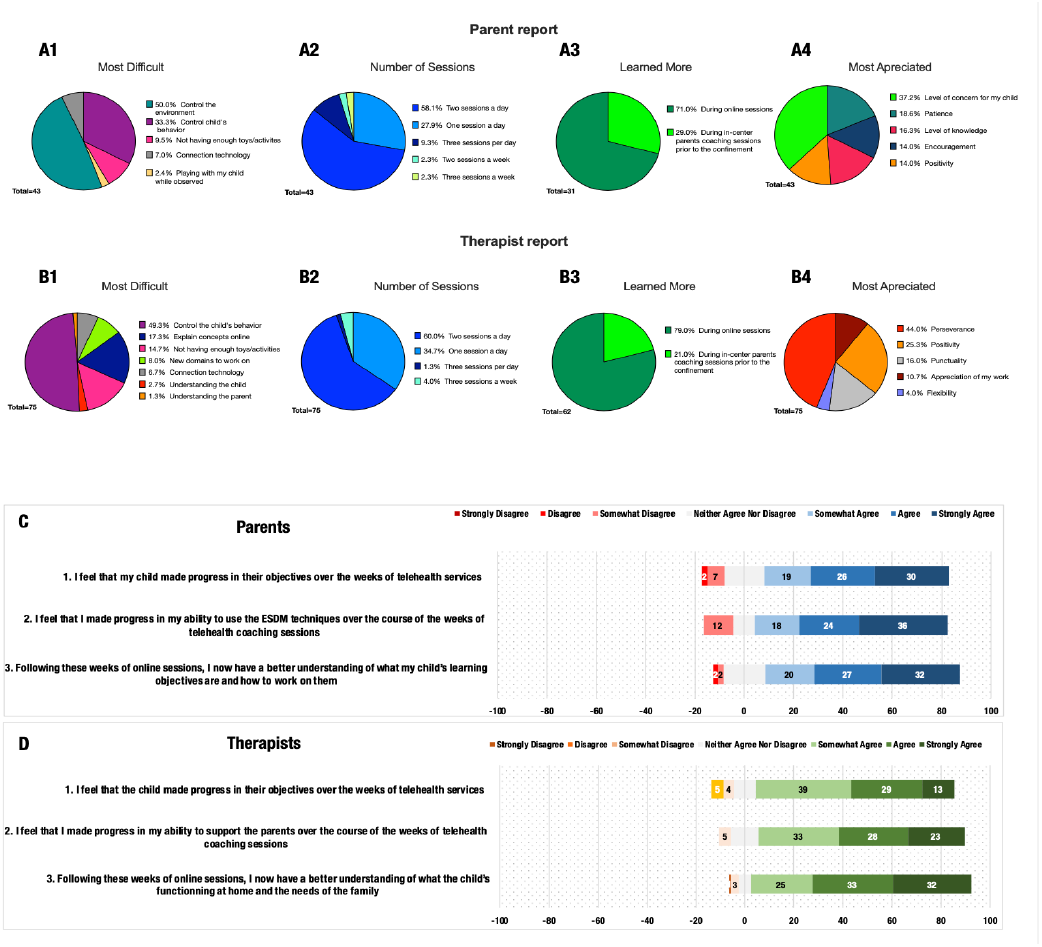
Satisfaction Questionnaire Administered Following the Final Remote Session *Note*. Panels A and B: multiple choice question answers for parents and therapists respectively; Panel C responses of the parents (n = 47); D therapists (n =45). The percentages of point responses in C and D are represented with the exception of the neutral point 4, *Neither agree nor disagree*, on the 7-point Likert scale.

#### 2. Parent Experience of Increased Frequency of PII Sessions

Nearly all parents (93%) felt that checking in daily with their child’s therapist made them feel supported in their efforts, and 86% felt that they learned more about ESDM techniques and were applying them more than before receiving the daily sessions (figure 3). Likewise, 84% of parents felt that they made progress in their ability to use the intervention techniques and 79% felt that the weeks of online daily sessions helped them to have a better understanding of their child’s learning objectives and how to work on them. Following the period of remote delivery, we asked the parents and therapists what, for them, would be the ideal frequency of sessions. To our surprise, 58% of parents and 60% of therapists felt that 2 sessions a day was ideal, while 28% of parents and 35% of therapists felt that one-session a day would have been better. None of the parents or therapists said they preferred the typical rate of 1x a week, however, two families and three therapists said they would have preferred only 2-3 sessions per week (figure 3, A2 & B2).

#### 3. Relationship Between Parent Reported Satisfaction and Socioeconomic Status

Questionnaire results were analyzed by three levels of family annual income (table 1). Overall, we did not find significant group differences in regards to participation or satisfaction of sessions, but noted a tendency for lower-income families to be more unanimously appreciative and positive about the services. All three groups reported that the sessions were very useful and helped their child progress, with less variance in the low-income group (figure 4, B). All groups reported high motivation to apply intervention strategies and valued the daily check-ins with the therapist. Mid-income families reported less technical issues compared to both low-income (U = 70·5; p = 0·07) and high-income families U = 48; p = 0·04) however, this last comparison did not withstand the Benjamini-Hochberg procedure for FDR control (Benjamini & Hochberg, 1995).

**Figure 4.**
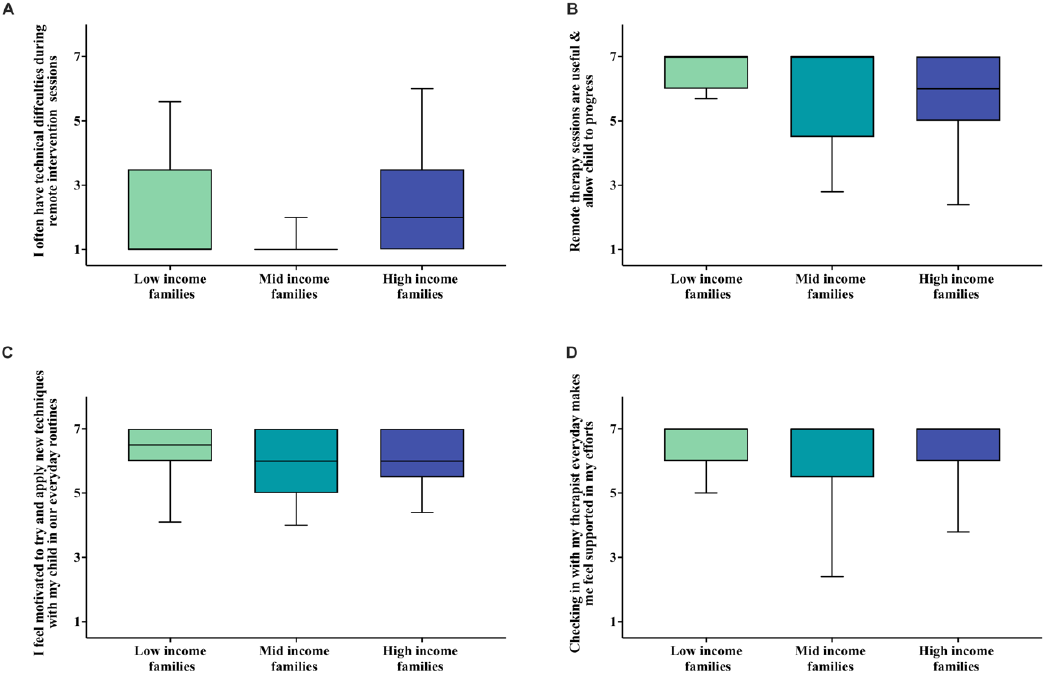
Parents’ Experience of Remote Sessions by Level of Family Annual Income *Note*. A: technology issues during remote sessions, B: perception of remote intervention usefulness; C: parents’ motivation to try new techniques; D: perceived importance of checking-in daily with the therapists of their child. Y-axis: 7-point Likert scale; X-axis: families represented by three income groups, as defined by the Swiss Federal Statistics Office guidelines.

#### 4. Comparison of parent and therapist satisfaction and perceptions

Overall, the therapists tended to be more hesitant about the telehealth format than the parents. In our assessment of parents’ comfort level with the new format, we found significant group differences between parents and therapists, with therapists perceiving the parents as less comfortable with the format than the parents actually were, both at Week 1(U = 649; p = 0·004, d = 0·6) and Week 3 (U = 986; p = 0·001, d = 0·6) (figure 5). Similarly, parents tended to perceive the therapists as more comfortable with the telehealth format than the therapists actually reported feeling. We found group differences both at Week 1 (U = 339; p < 0·001, d = 1·4) and Week 3 (U = 963; p < 0·001, d = 0·6). However, therapists showed a significantly higher estimation of their own comfort with the service delivery format at Week 3 compared to Week 1 (U = 1023; p < 0·001, d = 0.7).

**Figure 5.**
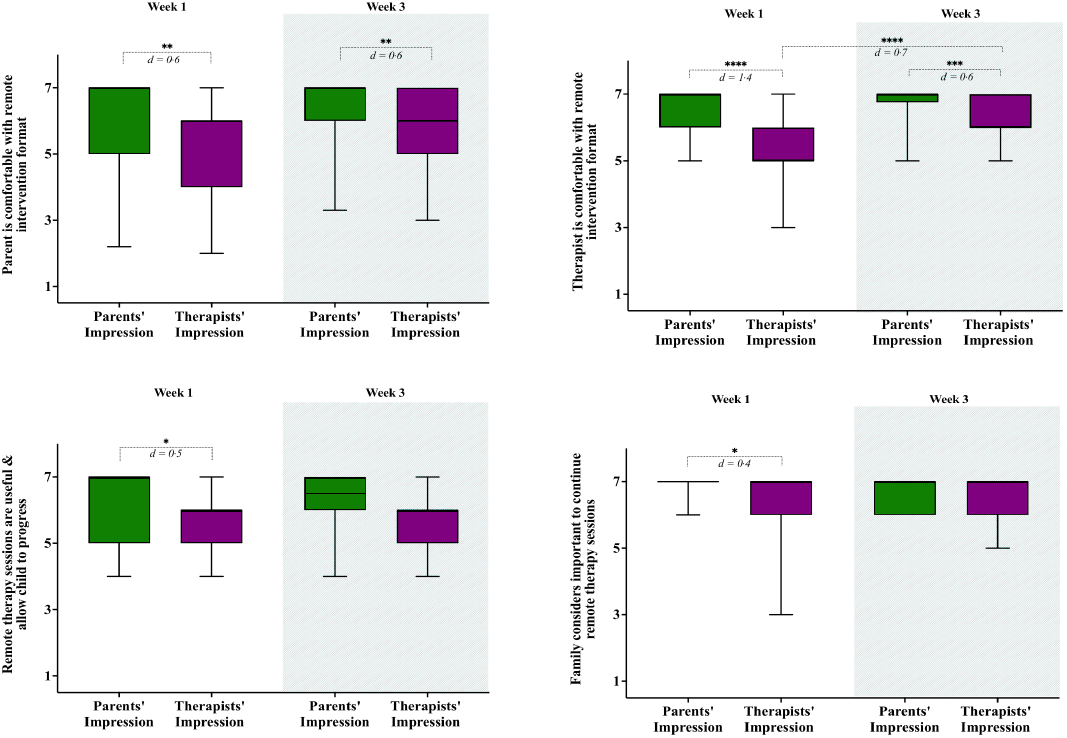
Comparison of Parent and Therapist Responses Impressions of Remote Sessions *Note*. Week 1 and Week 3 comparison of change in parents’ and therapists’ impressions with regards to: parental comfort with the new, remote format of intervention (upper left); therapists’ comfort with the remote format of intervention (upper right); perception of remote intervention usefulness (lower left) and family perception of importance to continue the online intervention (lower right). The effect size is expressed using Cohen’s d and significance level are defined as follows: * p < 0·05,** p < 0·01, *** p ≤ 0·001, **** p ≤ 0·0001.

We also found group differences for how useful the parents and therapists felt the daily sessions were. Parents perceived the sessions as being more useful than the therapists did, both at Week 1 (U = 687; p = 0·012, d = 0·5) and at Week 3 (U = 1145; p = 0·029, d = 0·4). However, the difference at Week 3 did not hold statistical significance after the Benjamini-Hochberg correction for multiple comparison (Benjamini & Hochberg, 1995). When we asked both parents and therapists whether they felt it was important to continue early intervention services remotely, we detected group differences at Week 1, with therapists having a slightly lower estimation of the importance to continue intervention during this period (U = 778; p = 0·020, d = 0·4). However, by the third week, both groups strongly agreed that it was important to continue the services remotely.

In the final questionnaire, we asked parents which characteristic they liked most about the therapist working with them (figure 3, A3). Thirty-seven percent of parents reported that the therapists “level of concern for my child” was what they appreciated most, with the others appreciating their “patience” (19%), “level of knowledge” (16%), “encouragement” (14%), and positivity (14%). Likewise, when the therapists were asked which qualities they most appreciated in the parents, 44% answered that it was the parents’ “perseverance”, while others felt it was the parents’ “positivity” (25%), “punctuality” (16%), “appreciation of my work” (11%), and “flexibility” (4%) that they liked most (figure 3, B3).

### Monitoring Child Developmental Progress

We aimed to measure the magnitude of change across different developmental domains assessed by the ESDM-CC following the intervention received during home-confinement. We compared the ESDM-CC global and sub-domain scores between three time points using Wilcoxon matched-pairs signed rank test. As depicted in figure 6, in all domains but expressive language we observed a pattern of continued significant improvement between all ESDM-CC evaluations, without any stagnation of progress related to the confinement period (between T2 and T3). For expressive language, the improvement observed between T2 and T3 did not reach significance level (p=0.06). Global scores showed significant change post (T3) compared to prior confinement (T2); Z = -2.71, p = 0.007, r = 0.36, with a slightly higher change observed in the period preceding the confinement (Z = -3.04, p = 0.002, r = 0.41). However, the change observed in three out of four assessed domains, namely Receptive Communication, Social Skills and Play was higher during the Confinement Period (T2-T3) (Z = -3.03, p = 0.002, r = 0.40, Z = -3.07, p = 0.002, r = 0.41, Z =

**Figure 6.**
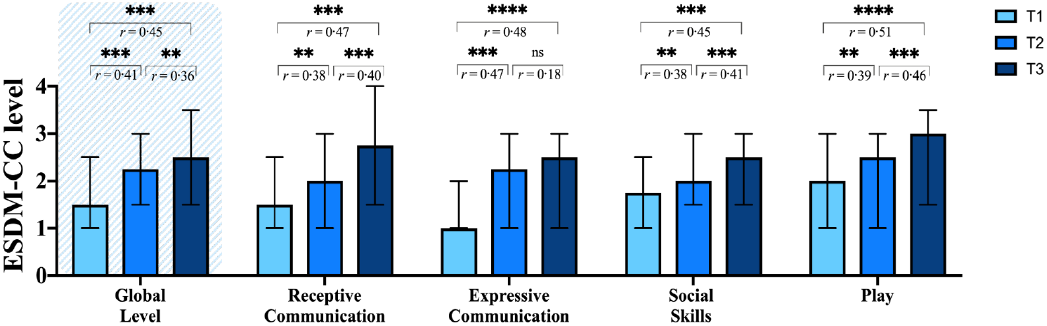
Change in ESDM-CC Levels at Three Time Points Note. Bar plots depicting the change in ESDM-CC levels at 3 time points: T1- ESDM-CC completed three-months prior to Pre-Confinement ESDM-CC, T2 - Pre-Confinement ESDM-CC, T3 Post-Confinement ESDM-CC. The magnitude of change is expressed in effect size r = Z/√ N (Rosenthal, 1991) and significance level are denoted as follows: ** p < 0·01, *** p ≤ 0·001, **** p ≤ 0·0001.

-3.45, p = 0.001, r = 0.46 respectively) compared to the pre-confinement change T1-T2 (Z = -2.82, p = 0.005, r = 0.38, Z = -2.81, p = 0.005, r = 0.38, Z = -2.94, p = 0.003, r = 0.39 respectively). This global effect might be driven by a lesser change evidenced in the Expressive Communication domain during T2-T3 (Z = -1.38, p = 0.17, r = 0.18) compared to the pre-confinement period (Z = -3.60, p < 0.001, r = 0.47).

## Discussion

### Remote Delivery of Parent-Implemented Intervention

Prior to Covid-19, remote delivery of autism services was mostly considered as a solution for families living far from service providers (Ellison et al., 2021; Parsons et al., 2017). The current study found that a telehealth format can be helpful to families regardless of where they live. For example, it allowed some of our working parents to join sessions during their breaks, while other parents appreciated the ease of having a coaching session at home while their younger child napped. For separated parents, it meant they could hear the same information without being physically in the same room. This supports findings from de Nocker and Toolan’s recent systematic review of telehealth autism services (de Nocker & Toolan, 2021) which suggest that increased accessibility, in particular for underserved communities, requires us to be open to more flexible intervention solutions (de Nocker & Toolan, 2021; Haine-Schlagel et al., 2020; Mirenda et al., 2022; Rogers et al., 2022; Vivanti et al., 2018).

Importantly, delivering services via videoconference allows professionals to work in the natural context of the child’s home, albeit virtually, instead of a clinic setting (Marino et al., 2020; Oono et al., 2013). Our programs had always provided in-person home visits, however, due to commuting time, only at an average rate of once-a-month. In the current study, both parents and therapists remarked on the benefits of having more frequent “in-home visits”, finding it helpful to have sessions in the environment in which the parent-child interaction typically takes place. Parents were able to show the therapists challenging behaviors as they occurred in the home, or prop their computer on the lunch table while getting help with their child’s feeding skills. This finding supports what other authors have identified as the importance of working on real-life situations outside of the clinical setting (Marino et al., 2020; Rogers et al., 2019; Vismara et al., 2018; Vivanti et al., 2018; Wetherby et al., 2014). Telehealth services in autism have, until recently, been thought of as an alternative to the preferable “in-person” meeting. The current study challenges this idea, suggesting the importance of a ‘hybrid’ PII model, where families benefit from both the live interaction of in-person coaching and the flexibility and reduced cost of remote intervention sessions.

### A Higher Frequency of Parent-Implemented Intervention Sessions

The telehealth format made possible an increased dosage of PII sessions, a key goal of which is to help parents acquire strategies to increase their child’s engagement and learning, and ultimately to encourage developmental progress (Oono et al., 2013; Shire et al., 2016). The idea of a higher dosage of parent coaching was explored in a recent study by Rogers and her colleagues (Rogers et al., 2019), where they compared weekly parent coaching with an enhanced program that involved bi-weekly sessions, demonstrating that caregivers who received the enhanced coaching made significantly greater gains in interaction skills with their child. Given the unique circumstances of the current study, we were not able to measure parent acquisition of skill, however, the majority of parents reported that the daily sessions helped them feel more confident in their ability to use and apply the intervention techniques. Parents also felt they learned more with the daily PII format than they had with the weekly, in-center coaching sessions received prior to the pandemic. This lends support to the idea that a higher frequency of parent coaching sessions is appreciated by caregivers, and may provide necessary support and motivation as they learn to implement the early intervention strategies (Parsons et al., 2017; Rogers et al., 2019; Wetherby et al., 2014).

Similarly, a 2014 randomized controlled study (Wetherby et al., 2014) demonstrated significantly greater improvements in child outcome in families receiving PII sessions two- or three-times a week compared to those receiving group parent-training once a week. In the current study, we were not able to include pre- and post-testing to measure child outcome, however, through analysis of our regular, quarterly monitoring of child developmental progress using the ESDM-CC, we observed a pattern of continued significant improvement across all developmental domains, except expressive language, where improvements were considerable but did not reach significance level. Although these findings do not allow us to draw conclusions on the efficacy of intensive PII, they do suggest that we should consider other forms of parent coaching than the once-a-week session, such as offering more frequent check-ins at the beginning of treatment to support and motivate parents as they learn the techniques and meeting more often thereafter, to encourage ongoing implementation. Further research is needed to determine whether a more intensive PII model leads to a more rapid acquisition and application of skills, and ultimately to child developmental progress.

### Remote Autism Intervention Services for a Diverse Community

The current study supports previous research showing that remote provision of autism intervention, including parent coaching, is not only feasible but well received by parents (Ellison et al., 2021; Ingersoll & Berger, 2015; Oono et al., 2013; Vismara et al., 2018). Unlike the majority of studies, however, our sample included comparable numbers of low-, mid- and high-income families, as well as parents with lower educational attainment, working parents, fathers, single-parents and families representing many different cultural groups (de Nocker & Toolan, 2021; Ingersoll & Berger, 2015; Oono et al., 2013; Rogers et al., 2019, 2022; Stahmer et al., 2019). This provided us with a more realistic picture of how telehealth autism services could work in our community setting. We found that nearly all of the families, regardless of background, were highly motivated to acquire the intervention tools, and appreciative of the regular support they received.

As Stahmer and her colleagues found in a 2019 study on improving access to autism services (Stahmer et al., 2019), income-based disparities, race and ethnicity have been shown to impact an autistic child’s access to care, with their families more likely to face barriers to participation. In the current study, home-confinement reduced some of these barriers, such as long working hours and transportation to and from therapy sessions. Nevertheless, our families of lower socioeconomic status tended to face more challenges, such as a smaller living space, fewer toys and materials, and lack of a computer and dependable internet connection. The fact that they were just as engaged in and satisfied with the sessions as their higher-income counterparts, suggests that families from diverse backgrounds regard their child’s programs very much as do those who, until recently, have been predominantly represented in the literature. However, for telehealth intervention to be more inclusive of traditionally underserved children, programs should ensure access to a computer, internet connection, toys and materials; and offer flexible scheduling to accommodate working-parents.

### Comparison of Parent and Therapist Experience

Responses to the questionnaires revealed significant differences, indicating that the therapists perceived the parents as being less at ease with the telehealth format than the parents reported feeling. Conversely, the parents tended to perceive their therapists as being more comfortable than the therapists reported feeling. These findings suggest that professionals may underestimate the readiness of families to participate in remote parent coaching, and may themselves come across as more confident than they actually feel. As with other studies looking at parent satisfaction with autism services, most of the parents in the current study reported that their therapist’s “level of concern for their child” was the characteristic they most appreciated, while therapists reported appreciating the parents’ “perseverance” and “positivity” (Estes et al., 2019; Makino et al., 2021; Mirenda et al., 2021).

### Limitations and Future Considerations

The main limitations of the current study stem from that which made it possible: the unexpected circumstance of providing autism intervention services during a sudden period of home-confinement. We had no opportunity to randomize groups, complete formal measures of child or parent outcome, control for ongoing therapist treatment fidelity, or trial our questionnaires. Despite the lack of formal metrics, the current study does offer a glimpse into community implementation of telehealth autism services, and demonstrates feasibility, even with limited preparation time. The beginning of the Covid-19 pandemic was, however, a period of great uncertainty, and this may have increased parents’ willingness to participate, in an effort to maintain continuity of their child’s program. It is also possible that the home-confinement condition meant that families had more free time to participate in parent coaching, however this is debatable, as the majority of parents in our study continued to work from home while also managing their children.

The level of satisfaction with the increased frequency of PII was high and pervasive. It may well have confounded the satisfaction reported with the telehealth format. This is somewhat of a moot point, however, since the increased frequency would not have been possible without the cost- and time-saving videoconference technology. The parents’ willingness to try remote intervention may have also been influenced by the fact that they already knew the therapists they would be meeting with. This fact may also have impacted their level of engagement and satisfaction with the sessions (Haine-Schlagel et al., 2020; Kronberg et al., 2021).

The Covid-19 pandemic disrupted our programs for children on the autism spectrum, forcing us to re-think our service delivery model and giving us the chance to experience more frequent interaction with their families. Although we have returned to a primarily in-person therapy model, we have increased our use of telehealth for parent support. This experience reminded us of the importance of parent-therapist partnership, and helped us imagine a future in which videoconferencing allows an increased frequency of parent coaching in the virtual home environment, opening up services to far more children in our community.

## Data Availability

The data used in this study are available upon a reasonable request to the 1st author of the study.

## Declarations

### Compliance with Ethical Standards

The relevant ethics committee regulating all work performed at the Faculty of Medicine of the Geneva University is the Commission Cantonale d’Ethique de la Recherche sur l’être humain; Regional Research Ethics Committee (CCER), Geneva, Switzerland. The CCER approved the current project (n° PB_2016-01880) on April 20, 2020. The methods used in the present study were performed in full accordance with the relevant guidelines and regulations of Geneva University.

### Consent

Informed consent was obtained from all individual participants included in the study, in accordance with protocols approved by the review board of the University of Geneva.

### Author contributions

Hilary Wood de Wilde wrote the original draft, Nada Kojovic completed data analysis and wrote the original draft for figures and table results; Supervision, review and editing by Marie Schaer and Edouard Gentaz. Céline Robertson, Catherine Karr, Leyla Ackman, Florence Caccia, Astrid Costes, Morgane Etienne, and Martina Franchini participated in the investigation, with Céline Robertson assisting with data curation. Project visualization, conceptualization and administration, as well as investigation, methodology, and data curation completed by Nada Kojovic, Marie Schaer and Hilary Wood de Wilde.

## References

American Psychiatric Association. (2013). Diagnostic and Statistical Manual of Mental Disorders (Fifth Edition). American Psychiatric Association. https://doi.org/10.1176/appi.books.9780890425596

Benjamini, Y., & Hochberg, Y. (1995). Controlling the False Discovery Rate: A Practical and Powerful Approach to Multiple Testing. Journal of the Royal Statistical Society. Series B (Methodological), 57(1), 289–300. JSTOR.

Crockett, J. L., Becraft, J. L., Phillips, S. T., Wakeman, M., & Cataldo, M. F. (2020). Rapid Conversion from Clinic to Telehealth Behavioral Services During the COVID-19 Pandemic. Behavior Analysis in Practice, 13(4), 725–735. https://doi.org/10.1007/s40617-020-00499-8

de Nocker, Y. L., & Toolan, C. K. (2021). Using Telehealth to Provide Interventions for Children with ASD: A Systematic Review. Review Journal of Autism and Developmental Disorders. https://doi.org/10.1007/s40489-021-00278-3

Ellison, K. S., Guidry, J., Picou, P., Adenuga, P., & Davis, T. E. (2021). Telehealth and Autism Prior to and in the Age of COVID-19: A Systematic and Critical Review of the Last Decade. Clinical Child and Family Psychology Review, 24(3), 599–630. https://doi.org/10.1007/s10567-021-00358-0

Estes, A., Swain, D. M., & MacDuffie, K. E. (2019). The effects of early autism intervention on parents and family adaptive functioning. Pediatric Medicine, 2, 21–21. https://doi.org/10.21037/pm.2019.05.05

Fuller, E. A., & Kaiser, A. P. (2020). The Effects of Early Intervention on Social Communication Outcomes for Children with Autism Spectrum Disorder: A Meta-analysis. Journal of Autism and Developmental Disorders, 50(5), 1683–1700. https://doi.org/10.1007/s10803-019-03927-z

Green, J., Charman, T., McConachie, H., Aldred, C., Slonims, V., Howlin, P., Le Couteur, A., Leadbitter, K., Hudry, K., Byford, S., Barrett, B., Temple, K., Macdonald, W., & Pickles, A. (2010). Parent-mediated communication-focused treatment in children with autism (PACT): A randomised controlled trial. The Lancet, 375(9732), 2152–2160. https://doi.org/10.1016/S0140-6736(10)60587-9

Haine-Schlagel, R., Rieth, S., Dickson, K. S., Brookman-Frazee, L., & Stahmer, A. (2020). Adapting parent engagement strategies for an evidence-based parent-mediated intervention for young children at risk for autism spectrum disorder. Journal of Community Psychology, 48(4), 1215–1237. https://doi.org/10.1002/jcop.22347

Ingersoll, B., & Berger, N. I. (2015). Parent Engagement With a Telehealth-Based Parent-Mediated Intervention Program for Children With Autism Spectrum Disorders: Predictors of Program Use and Parent Outcomes. Journal of Medical Internet Research, 17(10), e227. https://doi.org/10.2196/jmir.4913

Kronberg, J., Tierney, E., Wallisch, A., & Little, L. M. (2021). Early Intervention Service Delivery via Telehealth During COVID-19: A Research-Practice Partnership. International Journal of Telerehabilitation, 13(1). https://doi.org/10.5195/ijt.2021.6363

Makino, A., Hartman, L., King, G., Wong, P. Y., & Penner, M. (2021). Parent Experiences of Autism Spectrum Disorder Diagnosis: A Scoping Review. Review Journal of Autism and Developmental Disorders, 8(3), 267–284. https://doi.org/10.1007/s40489-021-00237-y

Marino, F., Chilà, P., Failla, C., Crimi, I., Minutoli, R., Puglisi, A., Arnao, A. A., Tartarisco, G., Ruta, L., Vagni, D., & Pioggia, G. (2020). Tele-Assisted Behavioral Intervention for Families with Children with Autism Spectrum Disorders: A Randomized Control Trial. Brain Sciences, 10(9), 649. https://doi.org/10.3390/brainsci10090649

Mirenda, P., Colozzo, P., Smith, V., Kroc, E., Kalynchuk, K., Rogers, S. J., & Ungar, W. J. (2022). A Randomized, Community-Based Feasibility Trial of Modified ESDM for Toddlers with Suspected Autism. Journal of Autism and Developmental Disorders. https://doi.org/10.1007/s10803-021-05390-1

Mirenda, P., Smith, V., Colozzo, P., Vismara, L. A., Ungar, W. J., & Kalynchuk, K. (2021). Training Coaches in Community Agencies to Support Parents of Children with Suspected Autism: Outcomes, Facilitators, and Barriers. Journal of Autism and Developmental Disorders. https://doi.org/10.1007/s10803-021-05363-4

Nahmias, A. S., Pellecchia, M., Stahmer, A. C., & Mandell, D. S. (2019). Effectiveness of community-based early intervention for children with autism spectrum disorder: A metaanalysis. Journal of Child Psychology and Psychiatry, 60(11), 1200–1209. https://doi.org/10.1111/jcpp.13073

Oono, I. P., Honey, E. J., & McConachie, H. (2013). Parent-mediated early intervention for young children with autism spectrum disorders (ASD). Cochrane Database of Systematic Reviews. https://doi.org/10.1002/14651858.CD009774.pub2

Parsons, D., Cordier, R., Vaz, S., & Lee, H. C. (2017). Parent-Mediated Intervention Training Delivered Remotely for Children With Autism Spectrum Disorder Living Outside of Urban Areas: Systematic Review. Journal of Medical Internet Research, 19(8), e198. https://doi.org/10.2196/jmir.6651

Robain, F., Franchini, M., Kojovic, N., Wood de Wilde, H., & Schaer, M. (2020). Predictors of Treatment Outcome in Preschoolers with Autism Spectrum Disorder: An Observational Study in the Greater Geneva Area, Switzerland. Journal of Autism and Developmental Disorders, 50(11), 3815–3830. https://doi.org/10.1007/s10803-020-04430-6

Rogers, S. J., & Dawson, G. (2010a). Early Start Denver Model curriculum cheklist for young children with autism. Guilford Press.

Rogers, S. J., & Dawson, G. (2010b). Early Start Denver Model for young children with autism: Promoting language, learning, and engagement. Guilford Press.

Rogers, S. J., Dawson, G., & Vismara, L. A. (2012). An early start for your child with autism: Using everyday activities to help kids connect, communicate, and learn. Guilford Press.

Rogers, S. J., Estes, A., Lord, C., Vismara, L., Winter, J., Fitzpatrick, A., Guo, M., & Dawson, G. (2012). Effects of a Brief Early Start Denver Model (ESDM)–Based Parent Intervention on Toddlers at Risk for Autism Spectrum Disorders: A Randomized Controlled Trial. Journal of the American Academy of Child & Adolescent Psychiatry, 51(10), 1052–1065. https://doi.org/10.1016/j.jaac.2012.08.003

Rogers, S. J., Estes, A., Vismara, L., Munson, J., Zierhut, C., Greenson, J., Dawson, G., Rocha, M., Sugar, C., Senturk, D., Whelan, F., & Talbott, M. (2019). Enhancing Low-Intensity Coaching in Parent Implemented Early Start Denver Model Intervention for Early Autism: A Randomized Comparison Treatment Trial. Journal of Autism and Developmental Disorders, 49(2), 632–646. https://doi.org/10.1007/s10803-018-3740-5

Rogers, S. J., Stahmer, A., Talbott, M., Young, G., Fuller, E., Pellecchia, M., Barber, A., & Griffith, E. (2022). Feasibility of delivering parent-implemented NDBI interventions in low-resource regions: A pilot randomized controlled study. Journal of Neurodevelopmental Disorders, 14(1), 3. https://doi.org/10.1186/s11689-021-09410-0

Rosenthal, R. (1991). Effect sizes: Pearson’s correlation, its display via the BESD, and alternative indices. American Psychologist, 46(10), 1086–1087. https://doi.org/10.1037/0003-066X.46.10.1086

Schreibman, L., Dawson, G., Stahmer, A. C., Landa, R., Rogers, S. J., McGee, G. G., Kasari, C., Ingersoll, B., Kaiser, A. P., Bruinsma, Y., McNerney, E., Wetherby, A., & Halladay, A. (2015). Naturalistic Developmental Behavioral Interventions: Empirically Validated Treatments for Autism Spectrum Disorder. Journal of Autism and Developmental Disorders, 45(8), 2411–2428. https://doi.org/10.1007/s10803-015-2407-8

Shire, S. Y., Gulsrud, A., & Kasari, C. (2016). Increasing Responsive Parent–Child Interactions and Joint Engagement: Comparing the Influence of Parent-Mediated Intervention and Parent Psychoeducation. Journal of Autism and Developmental Disorders, 46(5), 1737– 1747. https://doi.org/10.1007/s10803-016-2702-z

Stahmer, A. C., Vejnoska, S., Iadarola, S., Straiton, D., Segovia, F. R., Luelmo, P., Morgan, E. H., Lee, H. S., Javed, A., Bronstein, B., Hochheimer, S., Cho, E., Aranbarri, A., Mandell, D., Hassrick, E. M., Smith, T., & Kasari, C. (2019). Caregiver Voices: Cross-Cultural Input on Improving Access to Autism Services. Journal of Racial and Ethnic Health Disparities, 6(4), 752–773. https://doi.org/10.1007/s40615-019-00575-y

Vismara, L. A., McCormick, C. E. B., Wagner, A. L., Monlux, K., Nadhan, A., & Young, G. S. (2018). Telehealth Parent Training in the Early Start Denver Model: Results From a Randomized Controlled Study. Focus on Autism and Other Developmental Disabilities, 33(2), 67–79. https://doi.org/10.1177/1088357616651064

Vivanti, G., Kasari, C., Green, J., Mandell, D., Maye, M., & Hudry, K. (2018). Implementing and evaluating early intervention for children with autism: Where are the gaps and what should we do?: Gaps and priorities for intervention research. Autism Research, 11(1), 16–23. https://doi.org/10.1002/aur.1900

Wetherby, A. M., Guthrie, W., Woods, J., Schatschneider, C., Holland, R. D., Morgan, L., & Lord, C. (2014). Parent-Implemented Social Intervention for Toddlers With Autism: An RCT. Pediatrics, 134(6), 1084–1093. https://doi.org/10.1542/peds.2014-0757

Zwaigenbaum, L., Bauman, M. L., Choueiri, R., Kasari, C., Carter, A., Granpeesheh, D., Mailloux, Z., Smith Roley, S., Wagner, S., Fein, D., Pierce, K., Buie, T., Davis, P. A., Newschaffer, C., Robins, D., Wetherby, A., Stone, W. L., Yirmiya, N., Estes, A., … Natowicz, M. R. (2015). Early Intervention for Children With Autism Spectrum Disorder Under 3 Years of Age: Recommendations for Practice and Research. Pediatrics, 136(Supplement 1), S60. https://doi.org/10.1542/peds.2014-3667E

